# Global Analysis of an SEIRS Model for COVID-19 Capturing Saturated Incidence with Treatment Response

**DOI:** 10.1101/2020.05.15.20103630

**Authors:** David A. Oluyori, Helen O. Adebayo, Ángel G. C. Pérez

## Abstract

Sequel to V. A. Okhuese [Mathematical Predictions for COVID-19 as a Global Pandemic, *medRxiv*, 2020, https://doi.org/10.1101/2020.03.19.20038794], who studied the dynamics of COVID-19 using an SEIRUS model. We consider an SEIRS model capturing saturated incidence with treatment response. In this theoretical model, we assumed that the treatment response is proportional to the number of infected as long as the incidence cases are within the capacity of the healthcare system, after which the value becomes constant, when the number of confirmed cases exceed the carrying capacity of the available medical facilities. Thus, we obtain the reproduction number stating that when *R*_0_ is less than a critical value *R*, the disease-free equilibrium is globally asymptotically stable. Also, we studied the existence of the local and global stability of the disease-free and endemic equilibria and found that the kind of treatment response and inhibitory measures deployed in tackling the COVID-19 pandemic determines whether the disease will die out or become endemic.

## 1. Introduction

The novel coronavirus disease (COVID-19) was first confirmed in the Chinese city of Wuhan, late December 2019. The rapidity of its spread in many countries around the globe made the WHO declare it as a global pandemic and public health emergency, raising concerns that if countries with robust healthcare systems to detect and control disease outbreak are having challenges managing the disease, countries with weak healthcare system need to put adequate measures in place to contain the spread [1]. The coronavirus disease (COVID-19) caused by the Severe Acute Respiratory Syndrome Coronavirus-2 (SARS-CoV-2) presents clinical features which are similar to the diseases caused by other coronaviruses, Severe Acute Respiratory Syndrome (SARS) and Middle East Respiratory Syndrome such as lower respiratory illness with fever, dry cough, myalgia, shortness of breath etc. The Coronavirus disease is *“novel”* in the sense that, it is a new strain of zoonotic origin which has not been previously discovered to affect humans. Historically, the COVID-19 pandemic is a major human coronavirus epidemic in the last two decades aside SARS [2] and MERS [3, 4] respectively. The incubation period of COVID-19 is between 2–14 days with symptoms averagely between 5–7 days. Its basic reproduction number is averaged 2.2 [5] and even more ranging from 1.4 – 6.5 in [6]. Globally as at May 5^th^, 2020, there are 3,646,304 confirmed cases, 1,200,296 recovered and 252,425 deaths.

The *SARS-CoV-2* is an enveloped positive-sense stranded RNA virus (*ssRNA*) (Subgenus*: Sarbecovirus, Orthocoronavirinae* subfamily), consisting of 29,903 nucleotides and two untranslated sequences of 254 and 229 nucleotides at the 5’ and 3’-ends respectively (GenBank No. MN 908947) [7]. The coronaviruses (*CoV*) are subdivided into 4 genera *α–, β–, γ–, δ–*coronaviruses. The *α–*and *β–* CoV infect mammals while the *γ–*and *δ–CoV* infect birds. Previously, 6-*CoVs* have been identified as human susceptible virus, which are *α–CoVs*, *HCoV –*229E, *HCoV –NL*63, *β –CoVs*, *HCoV –HKUI* and *HCoV –OC*43. Specifically, SARS-CoV-2 is of the *β–*type, it enters the human epithelial via the binding of the viral spike protein (*S–Protein*) to the human Angiotensin Converting Enzyme 2 (ACE-2), which is a functional receptor involved in arterial blood pressure control and fibrotic response to damage, using it as its binding site. [8, 9]. Basically, there are 3 main transmission routes for the disease, which are: *Droplets transmission*, which occurs when respiratory droplets produced by infected persons when they cough or sneeze in close proximity with other people are inhaled or ingested; *Contact transmission*, which occur when people touch infected surfaces and subsequently touch their mouth, nose or eyes; *Aerosol transmission*, which occurs when respiratory droplets mix with the air, forming aerosols are inhaled in high doses, which are potent enough to cause infection [11].

### 1.1. Models for incidence and treatment parameters

According to [20], *Incidence* is the rate at which the susceptible become infectious. Thus, if the unit of time is days, then the incidence is the number of new infections per day. The classical Kermack-McKendrick model in [21] proposed a *Bilinear Incidence* rate for simple mass action denoted as *h*(*I*)= *βSI*, where *S* and *I* denote the susceptible and Infected respectively and *β* denote the probability of transmission per contact. Next, we have *Standard Incidence*, which is denoted as 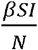, where *N* is the total population size, *β* is the average number of effective contacts per unit time of an infective with the susceptible (daily contact rate), since 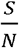 is the susceptible fraction of the population, 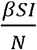 is the average number of infection transmissions by all infectives. According to literature, the bilinear and standard incidences have been extensively studied by various authors in [33-36] and others. Another kind of incidence that is of interest to our work is the *Saturation incidence*. In 1973, sequel to the study of cholera epidemic which occurred in Bari, Capasso and Serio [22] introduced the saturated incidence denoted as *h*(*I*)*S* into epidemic models, where *h*(*I*) tends to a saturation level when *I* becomes large, i.e.,

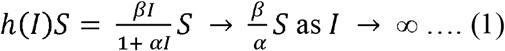

where *βI* denotes the infection force of the disease and 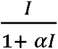 measures the inhibitory effect from the behavioural change of the susceptible when their number of incidence increases or from the crowding effect of the infective due to unrestrained contact using suitable parameters. Esteva and Matias [20] studied a model for disease transmitted by vector with saturating incidence such that the model assumes a saturating effect in the incidence due to the response of the vector to change in the susceptible and infected densities. The saturation incidence seems more realistic than the bilinear incidence due to the inclusion of behavioural change and crowding effect of the infective. In the face of the current realities from COVID-19, it is evident that we have high saturation incidence in which useful strategies need to be deployed to contain the spread through various interventions such as good hygiene, physical/social distancing, partial/total lockdown, travel/public gathering ban, good treatments, contact tracing, pool testing, etc, can help to reduce the high rate of secondary infections as stipulated by the WHO guidelines.

It is a general assumption in classical epidemic models that treatment rate of infection is assumed to be proportional to the number of the infective individuals and the recovery rate depends on the medical resources available such as test kits, drugs, isolation centres, ventilators, availability of trained medical personnel, efficiency of treatment. WHO situation reports from many nations have shown how stretched the healthcare systems of countries have been with its attendant high morbidity. Therefore, it is important for countries with increased cases to adopt suitable treatment functions. Wang and Ruan [23] introduced a constant treatment in SIR models as follows:

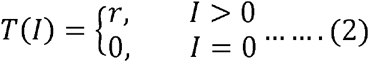

where *r* is a positive constant and *I* is the number of infected individuals.

Recently, Wang [24] considered a piecewise linear treatment function defined as:

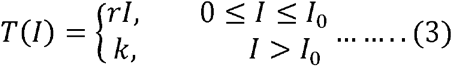

where *r=rI*_0_, *r and I*_0_ are positive constants.

The first condition in (3) explains the proportionality of the treatment response to the number of the infectious people when the number of infectious is less than or equal to a fixed value *I*_0_, the second typifies an endemic situation (*I > I*_0_), where the number of the infectious has increased to a saturation point where the available medical facilities are stretched beyond capacity and death toll rises in an unprecedented manner. Therefore, in many disease outbreaks there are different kinds of delays when they spread, such as latent period delay before symptoms surfaces and immunity period delay after recovery. Zhou [25] studied an SIR epidemic model with treatment function 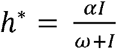. Zhang et’al [26] modified the model in [25], with saturated incidence rate, 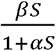 using the same treatment function. In [27] Agrawal et’al modified the work of [26] considering an SEIRS epidemic model with saturated incidence and treatment rate. Badole et’al in [28] taking some cue from [27] studied the global dynamics of an SEIR model with saturated incidence under treatment. Various authors have considered saturated incidence and treatment to study the stability and bifurcation of different dynamic systems in [24, 34, 37-39].

### 1.2. Existing compartmental models for COVID-19

Since the outbreak, many mathematical have appeared in an attempt to assess the dynamics of the COVID-19 epidemic. The first models were dynamic mechanistic models aimed at estimating of the basic reproduction number *R*_0_ [12, 13, 14, 15], also simple exponential growth models [16, 17]. Other compartmental epidemiological models such as SIRD, SEIR, SEIRD and SEIRUS [21, 19, 18, 10] has been proposed to estimate other epidemiological parameters such as the transmission rate, local and global stability of the disease-free and endemic equilibria to provide insights for forecasting purposes.

Recently, [10] considered an SEIRUS (Susceptible-Exposed-Infected-Recovered-Undetectable-Susceptible) model for COVID-19, where it was predicted that with strict adherence to the guidelines of the WHO on observatory and treatment procedures, the pandemic will soon die out. Based on the motivation from [10, 24, 27, 28, 29], we present an SEIRS (Susceptible-Exposed-Infected-Recovered-Susceptible) model with saturated incidence and treatment functions which prescribes inhibitory measures such as personal hygiene, wearing of face mask, travel/public gathering ban, partial or total lockdown etc and rapid responses such as public enlightenment, pool testing, increased medical facilities and trained medical personnel etc, as potent means of slowing down the spread of COVID-19.

## 2. Model description

The model can be described as follows:

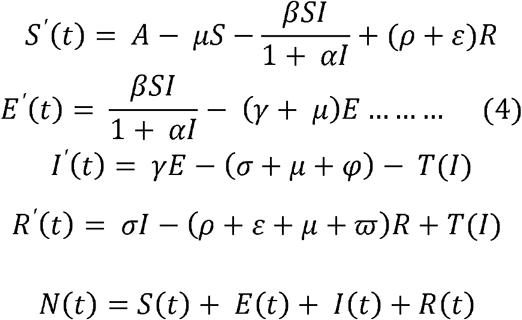

Here, *S*(*t*) is the number of susceptible per unit time, *E*(*t*) is the number of the exposed per unit time, *I*(*t*) is the number of the infectious per unit time, *R*(*t*) is the number of the recovered per unit time, *A* is the recruitment rate of the population, *μ* is the natural death rate of the population per time, *ρ* is the recovery rate, *α* is the saturation parameter that measures the inhibitory effect, *β* is the rate of transmission, ε is the proportion of the removed population that is been observed and will subsequently move to the susceptible, *γ* is the rate of developing infection/incidence rate. *φ* is the disease-induced death rate of the infected population not quarantined, *ϖ* is the fraction of the removed population under observation (the undetected) before moving to the susceptible class, 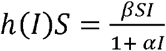 is the saturation incidence parameter, 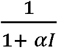 is the inhibitory parameter and *T*(*I*) is the treatment response as defined in (3). The flow diagram of the model can be seen in Figure 1.

**Figure 1.**
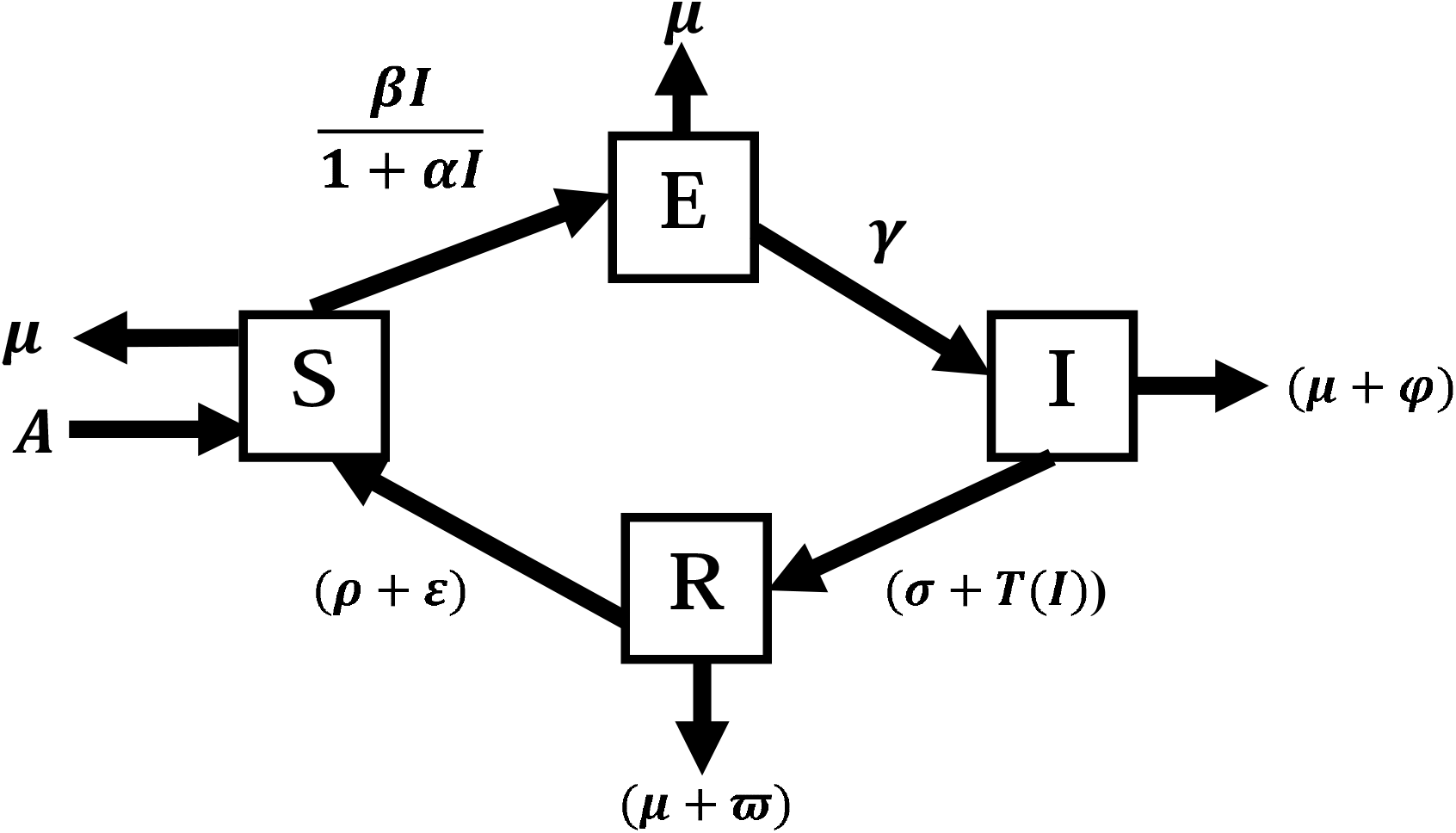
Schematic diagram of the model.

It follows from (4) that

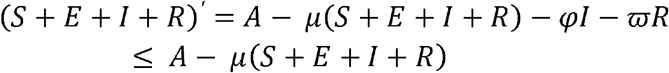

Then 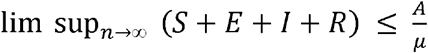. The feasible region for system (4) is

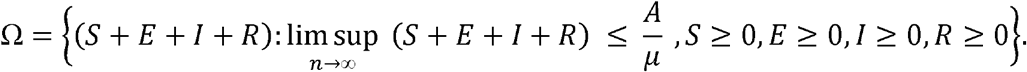

Thus, it naturally follows that the region Ω is positively invariant with respect to system (4). Hence the system is mathematically and epidemiological well posed in Ω.

## 3. Dynamics near the disease-free equilibrium

We will now study the local dynamics of system (4) near the disease-free equilibrium. Thus, we will focus here on the case when 0 *≤ I ≤ I*, that is, when *T*(*I*) = *rI*. In that case, system (4) becomes

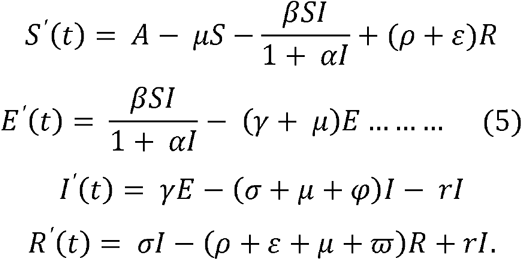

By simple calculation from the system (5), we obtain the equilibrium state where *S′*(*t*)= *E′*(*t*)*=I′*(*t*)*=R′*(*t*)=0 (i.e. the LHS vanishes). Thus the steady state of system (5) satisfies the following algebraic system of equations:

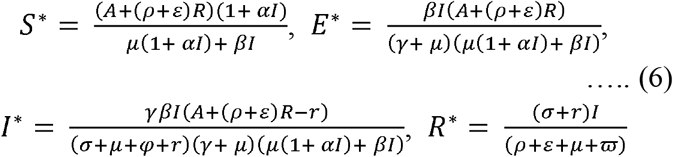

At the disease-free equilibrium (DFE), when no disease outbreak occurs, no one is in the exposed or infected class and as such no one is in the recovered class. Therefore, *E = I* = 0. On substitution, the algebraic system in (6) reduces to 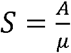.

Thus, we conclude the following result.

### Lemma 3.1

The system (5) always has a disease-free equilibrium point 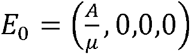.

### 3.1. Basic reproduction number

The basic reproduction number *R*_0_ measures the probability of an infectious disease spreading through a population or becoming extinct after some time. This threshold characteristic of *R*_0_ helps epidemiologist to make the following assumptions: (a) If *R*_0_ *<1*, the infection will die out with time and (b) If *R*_0_ *> 1*, the disease will be endemic in the population.

Next, we find the reproduction number, *R*_0_ of the system (4) by obtaining the Jacobian of the system and using the Next Generation Matrix due to Van den Driessche and Watmough [32].

The Jacobian of system (4) when 0 ≤ *I* ≤ *I*_0_ is given by

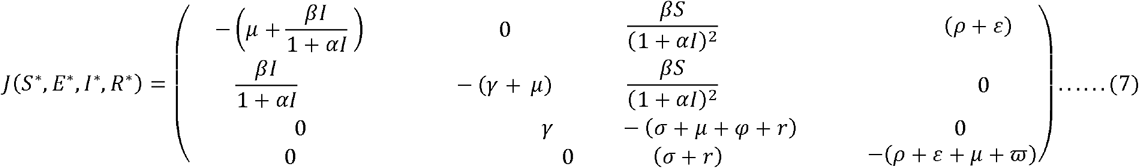

Applying the disease-free state condition 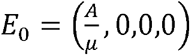 in the Jacobian matrix, we have

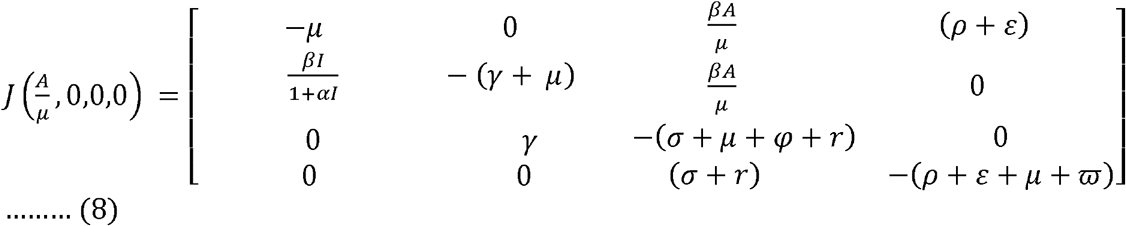

Using the next generation matrices, it is clear that the reproduction number *R*_0_ is the spectral radius of the next generation matrix derived from the exposed and infected class, i.e.,

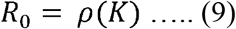

where *ρ*(*K*) is the spectral radius and *K=FV*^−1^ is the next generation matrix. *F* is derived from the exposed and infected class and *V* are the remaining terms after *F* is taken.

Thus

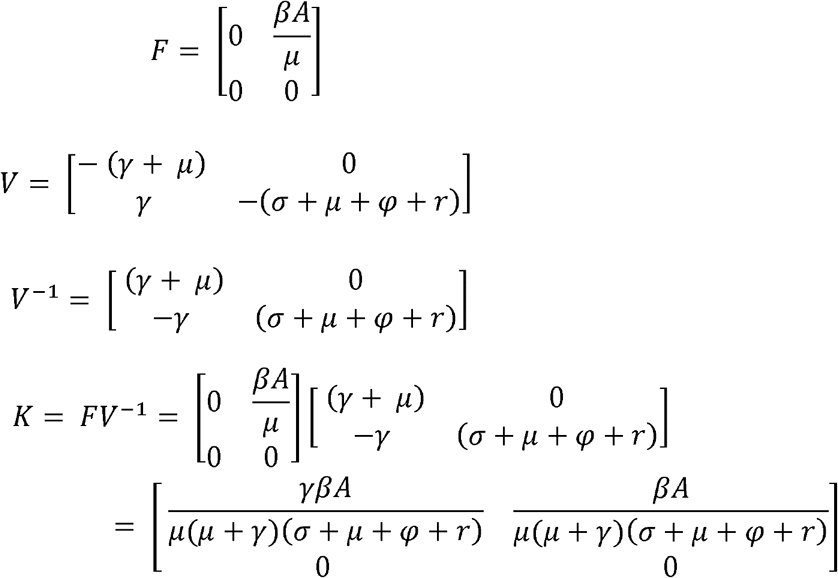

*K* is the next generation matrix of the system (4).

The spectral radius is

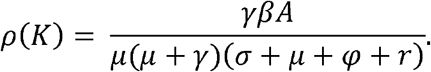

Hence the basic reproduction number *R*_0_ of system (4) is

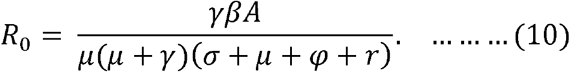

### 3.2. Local stability analysis of the disease-free equilibrium

We examined the local stability of the DFE by the analysis of the eigenvalues of the Jacobian matrices of (4) at the equilibrium using the *Routh-Hurwitz Criterion*.

#### Theorem 3.2

The disease-free equilibrium (*E*^0^)is

(a) Locally asymptotically stable if *R*_0_ *<* 1.

(b) Unstable if *R*_0_ *> 1*.

##### Proof

The Jacobian matrix of the system (4) at the disease-free equilibrium is

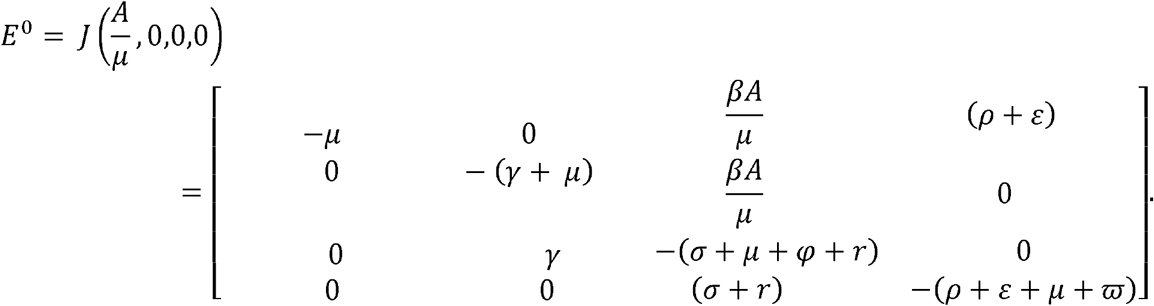

The characteristic equation of the system (5) at *E*^0^ is

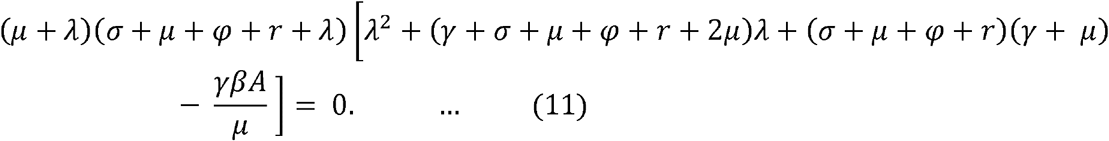

By (11) it is clear that *λ*_1_ *=−μ* and *λ*_2_*=−*(*σ + μ + φ + r*) are two roots of (11). The other roots of (11) are determined by the equation

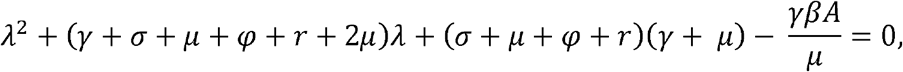

which has negative roots if and only if 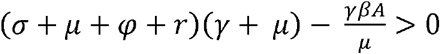, which is equivalent to the reproduction number *R*_0_ being less than one. This implies that the disease-free equilibrium *E*^0^ is locally asymptotically stable when *R*_0_ *<* 1 and unstable when *R*_0_ *>* 1.

## 4. Existence of endemic equilibria

In this section, we consider the endemic equilibria of system (4). An endemic equilibrium of (4) satisfies the system

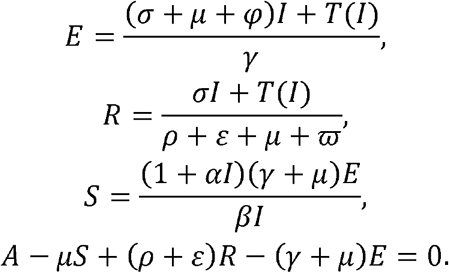

Suppose that *P^*^*(*S^*^, E^*^, I^*^, R^*^*) is an endemic equilibrium of (4). If 0 *< I^*^ ≤ I*_0_, then *P^*^* satisfies

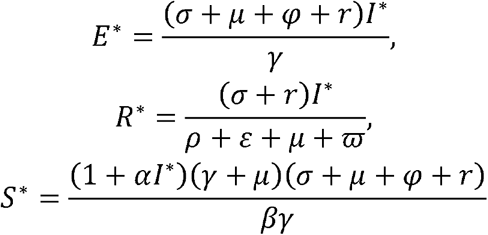

and

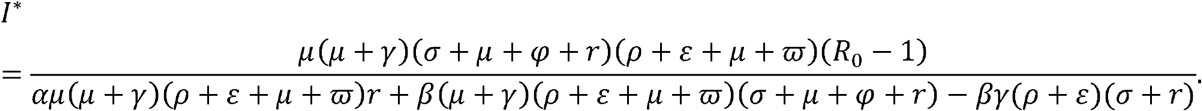

The denominator of the expression for *I^*^* is always positive, while the numerator is positive if and only if *R*_0_ *> 1*. The condition *I^*^≤ I*_0_ is equivalent to 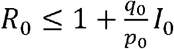, where

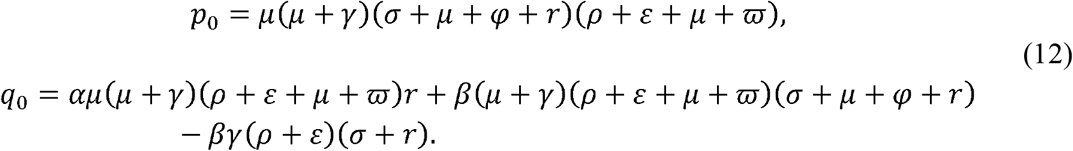

Therefore, we can conclude the following result.

### Theorem 4.1

*P^*^*(*S^*^, E^*^, I^*^, R^*^*) is an endemic equilibrium of (4) if and only if

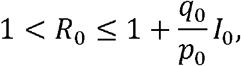

where *p*_0_ and *q*_0_ are given by (12).

Next, we study the endemic equilibria of (4) when *I > I*_0_ and hence *T*(*I) = r*.

An endemic equilibrium (*S, E, I, R*) with *I > I*_0_ satisfies

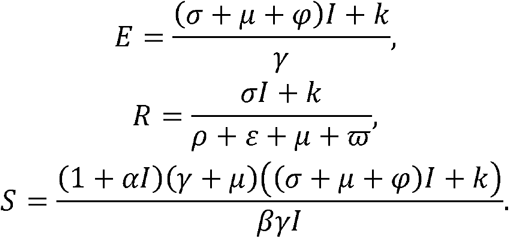

Substituting these expressions in the equation *A − μS +* (*ρ + ε)R−*(*γ + μ) E* = 0, we obtain the quadratic equation

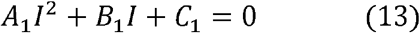

where

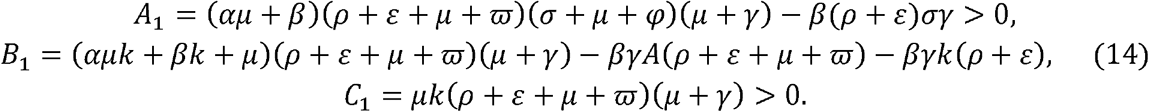

If *B*_1_ ≥ 0, it is clear that (13) does not have any positive solutions. Consider the case when *B*_1_*<*0. Let 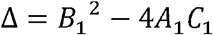. The solutions are given by

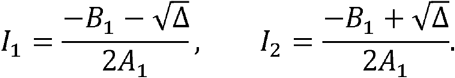

These solutions are positive and distinct only when Δ *>* 0. The two solutions coalesce into a double positive root when Δ = 0. Lastly, (13) has no real solutions when Δ *<* 0. Hence, we obtain the following result.

### Theorem 4.2

Let *A*_1_, *B*_1_ and *C*_1_ be given by (14). Define 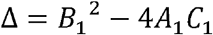,

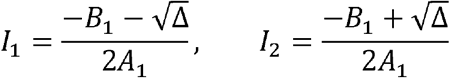

and

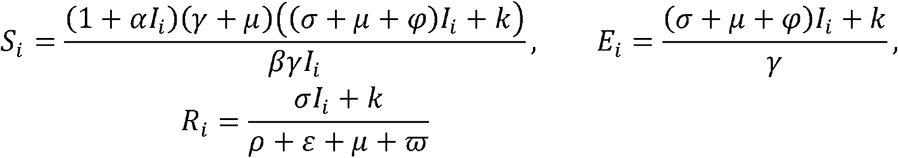

for *i*=1, 2. Then *P*_i_(*S*_i_ *E*_i_, *I*_i_, *R*_i_) is an endemic equilibrium of (4) if and only if *B*_1_ *<* 0, Δ ≥ 0 and *I_i_* > *I*_0_. Moreover, *P*_1_ and *P*_2_ coalesce into a single endemic equilibrium when *B*_1_ *<* 0, Δ = 0 and *I*_1_ = *I*_2_ *> I*_0_.

## 5. Global stability of the disease-free equilibrium

In this section, we analyze the global stability of the disease-free equilibrium for system (4) using the method of Lyapunov functions.

### Theorem 5.1

Let

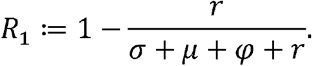

If *R*_0_ *<R*_1_, then the disease-free state equilibrium *E*^0^ is globally asymptotically stable.

#### Proof

From the system of equations in (5), we have 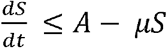. A solution of the equation 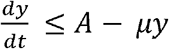 is a maximal solution of *S*(*t*). We recall that 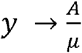 as 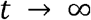. By applying the comparison theorem, we obtain 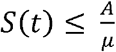, also from the set

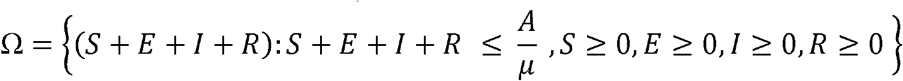

we have 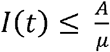.

Define the Lyapunov function

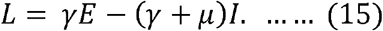

From 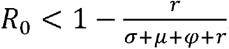 we have *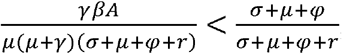*, and then *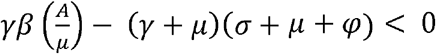*

We have thus *L*′ = *γE*′*−*(*γ + μ)I*′, that is,

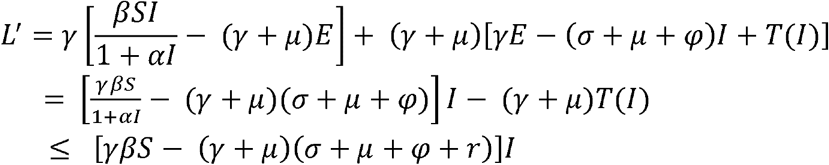

Recall that 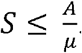. Therefore,

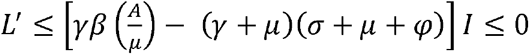

and *L*′ = 0 if and only if *I* = 0. Thus, the largest compact invariant set in *{*(*S, E, I, R)∈ Ω, L′* = 0} is the singleton *E*^0^. Therefore, by Lasalle-Lyapunov theorem, every solution that starts in *Ω* approaches *E*^0^ as *t* → *∞* and the proof is complete. ■

## 6. Local stability of endemic equilibria

We consider the local stability of the endemic equilibrium point *P*^*^(*S*^*^, *E*^*^, *I*^*^, *R*^*^) by analyzing the eigenvalues of the Jacobian matrices of (4) at the endemic equilibrium point using the *Routh-Hurwitz Criterion*.

### Theorem 6.1

Let *P*^*^(*S*^*^, *E*^*^, *I*^*^, *R*^*^) be an endemic equilibrium of (4) with *I^*^≤ I*_0_. Then *P^*^* is locally asymptotically stable if and only if *a*_3_ *>* 0, *a*_4_ *>* 0 and 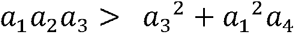, where *a*_1_, *a*_2_, *a*_3_ and *a*_4_ are given in (17).

#### Proof

When *I^*^≤ I*_0_, system (4) can be written as (5). The Jacobian of system (5) at the endemic state *P*^*^ is

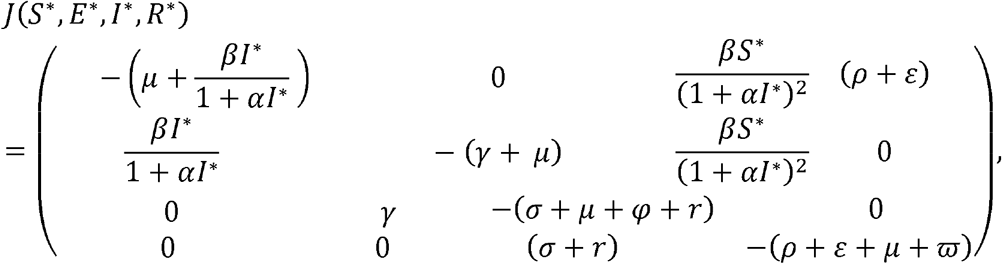

from which we obtain the characteristic equation

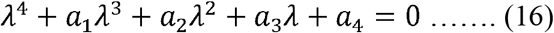

where *a*_1_, *a*_2_, *a*_3_, and *a*_4_ are as defined below:

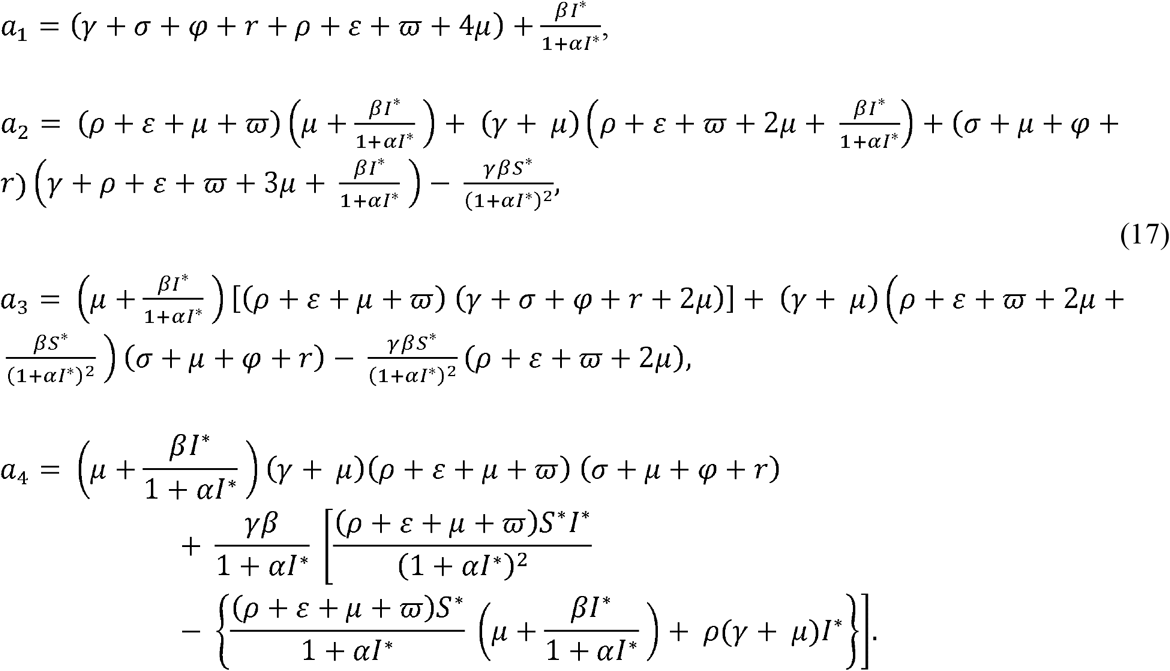

Notice that *a*_1_ is always positive. Thus, by the Routh-Hurwitz criterion, we have that the endemic equilibrium *P^*^*(*S*^*^, *E*^*^, *I*^*^, *R*^*^) of (4) is locally asymptotically stable if and only if *a*_3_*>*0, *a*_4_*>*0 and 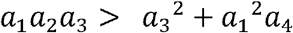. Hence, the theorem follows.■

Next, we discuss the local stability for the second case of the treatment function, that is, when *T*(*I) = k*, which corresponds to an equilibrium with *I^*^≤ I*_0_. In this case, we have the system

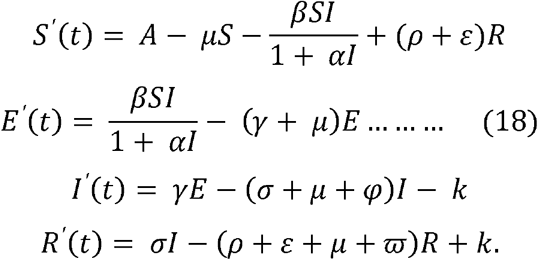

### Theorem 6.2

Let *P*^*^(*S*^*^, *E*^*^, *I*^*^, *R*^*^) be an endemic equilibrium of (4) with *I^*^> I*_0_. Then *P^*^* is locally asymptotically stable if and only if *b*_1_*>*0, *b*_4_*>*0 and 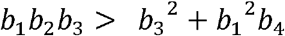, where *b*_1_, *b*_2_, *b*_3_ and *b*_4_ are given in (21).

#### Proof

The Jacobian matrix of system (18) at *P^*^* is

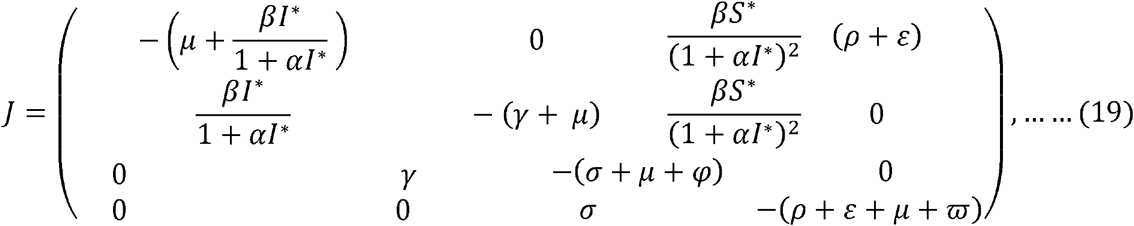

from which we obtain the characteristic equation

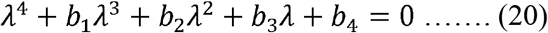

where *b*_1_, *b*_2_, *b*_3_, and *b*_4_ are as defined below:

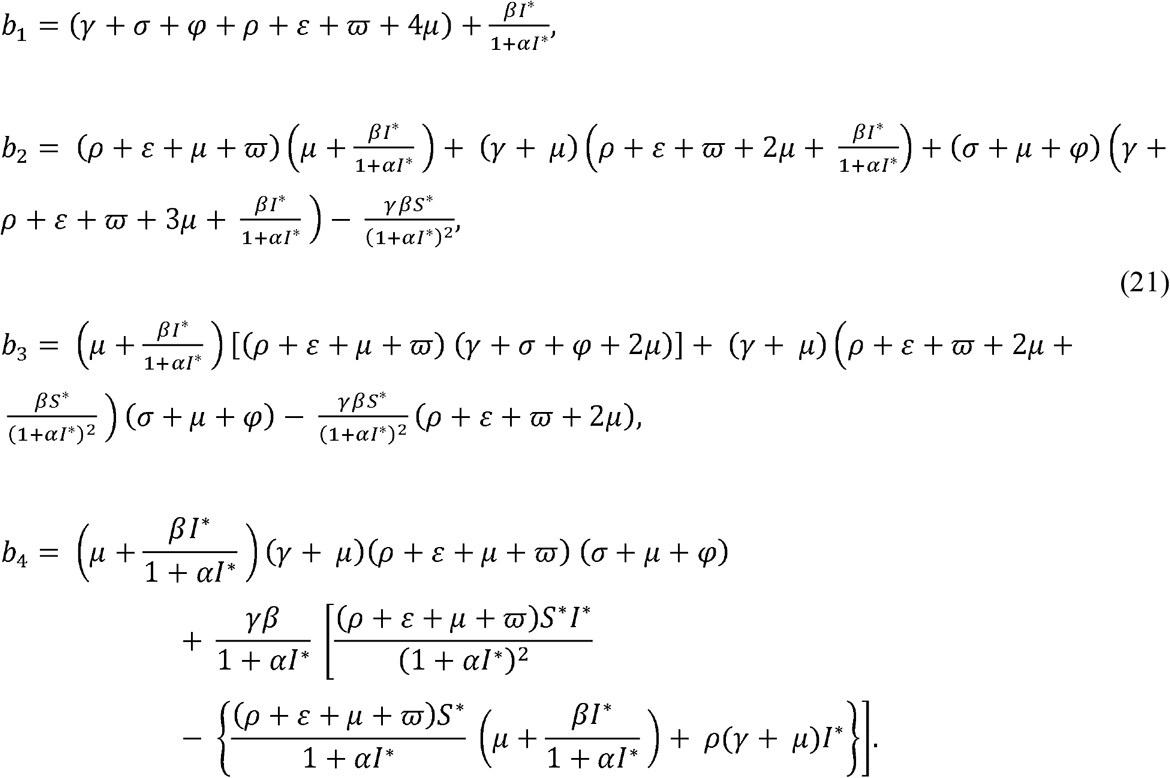

It is clear that *b*_1_ is always positive. Thus, by the Routh-Hurwitz criterion, we have that the endemic equilibrium *P*(*S*^*^, *E*^*^, *I*^*^, *R*^*^) of (4) is locally asymptotically stable if and only if *b*_3_*>*0, *b*_4_*>*0 and 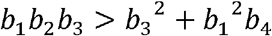. Hence, the theorem follows. ■

## 7. Global stability of endemic equilibria

In this following, we analyze the global stability of the endemic equilibrium when *I^*^≤ I*_0_. To do this, we reduce the system of equations in (5) using *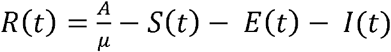* to eliminate the *R*(*t*) component from the first equation of system (5) to obtain a three-dimensional system given below:

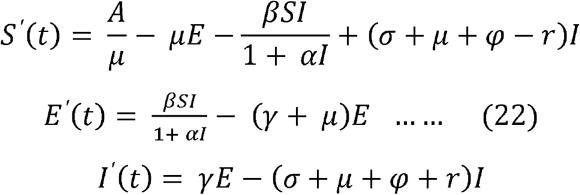

with initial conditions *S ≥* 0, *E ≥* 0*,I ≥* 0.

We consider the geometric approach due to Li and Muldowney [31] to obtain the global stability of the endemic equilibrium and find the sufficient conditions for which the endemic equilibrium is globally asymptotically stable. We describe the geometric approach method as follows. We consider

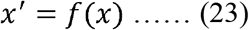

where *f: D* → *R^n^, D* ∩ *R^n^* is a simply connected open set and *f ∈ C*^1^(*D)*.

Let *x*^*^ be an equilibrium of the equation (23), i.e., *f*(*x*^*^)=0. Assume that the following hypotheses hold.

*(YI):* There exists a compact absorbing set *K ⊂ D*.

*(Y2):* Equation (23) has a unique equilibrium *x*^*^ *∈ D*.

The basic idea in this method is that if the equilibrium *x*^*^ is locally stable, then the stability will hold provided the conditions in (*Y1)* and (*Y2)* are satisfied and no non-constant periodic solution of (23) exists. Thus, the sufficient conditions on [ capable of precluding the existence of such solutions must be found.

Suppose that the conditions (*Y1)* and (*Y2)* is satisfied. Assume that (23) satisfies a Bendixson criterion that is robust under *C*^1^ local perturbations of *f* at all non-equilibrium non-wandering points for (23). Then *x*^*^ is globally stable in *D* provided it is stable. Let *P*(*x)* be an 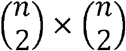 matrix valued function that is *C*^1^ on *D* and consider

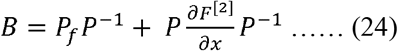

where the matrix *P_f_* is

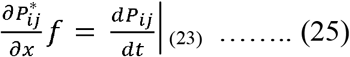

and the matrix *J*^[2]^ is the second additive compound matrix of the Jacobian matrix *J*, that is, *J*(*x*) = *Df*(*x)*. Generally speaking, for an *n×n* matrix *J*=(*J_ij_*), *J*^[2]^ is an 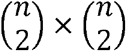 matrix and in the special case *n*=3 one has

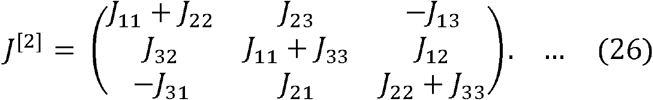

Consider the Lozinskiĭ measure *μ* of *B* with respect of a vector norm ║·║ in 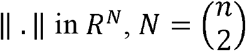, defined by

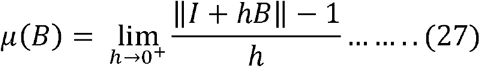

Suppose that (*Y*1) and (*Y*2) hold and consider the condition

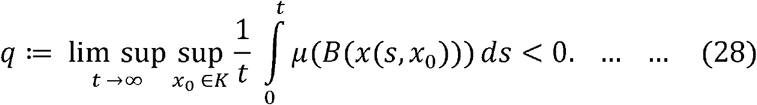

It is shown in [31] that, if *D* is simply connected, the condition *q<*0 rules out the presence of any orbit that gives rise to a simple closed rectifiable curve that is invariant for (23), such as periodic orbits, homoclinic orbits and heteroclinic cycles. Moreover, it is robust under *C*^1^ local perturbations of *f* near any non-equilibrium point that is non-wandering. In particular, the following global stability result is proved in [31].

### Lemma 7.1

Assume that *D* is simply connected and that the assumptions (*Y1)* and (*Y2)* is satisified. Then the unique equilibrium *x^*^* of (23) is globally stable in *D* if *q<*0. *■*

Having established the above, we study the global stability of the endemic equilibrium *P*^*^ and obtain the following result.

### Theorem 7.2

Let *P*^*^(*S*^*^, *E*^*^, *I*^*^, *R*^*^) be an endemic equilibrium of (4) with *I*^*^ ≤ *I*_0_. If *μ*−γ *>* 0 then the endemic equilibrium *P*^*^ is globally stable.

#### Proof

Since *I*^*^ ≤ *I*_0_, (*S*^*^, *E*^*^, *I*^*^) is an equilibrium for system (22). The Jacobian matrix of system (22) is given as

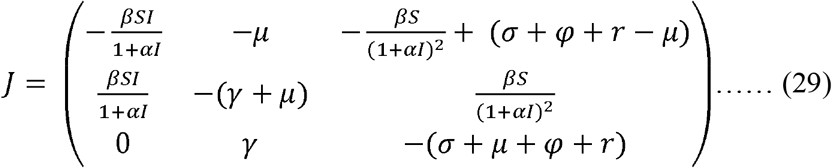

and its second additive matrix is

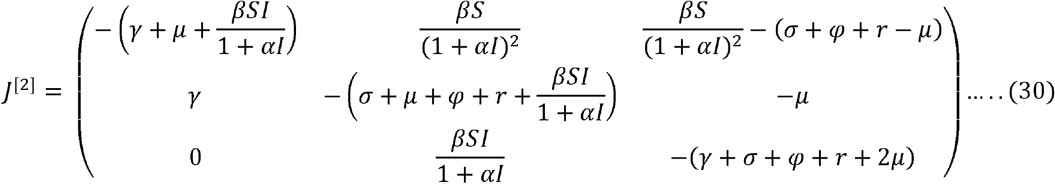

Choose the function 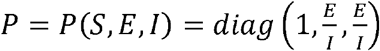; then 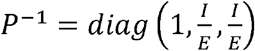 and

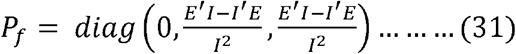

Then we have

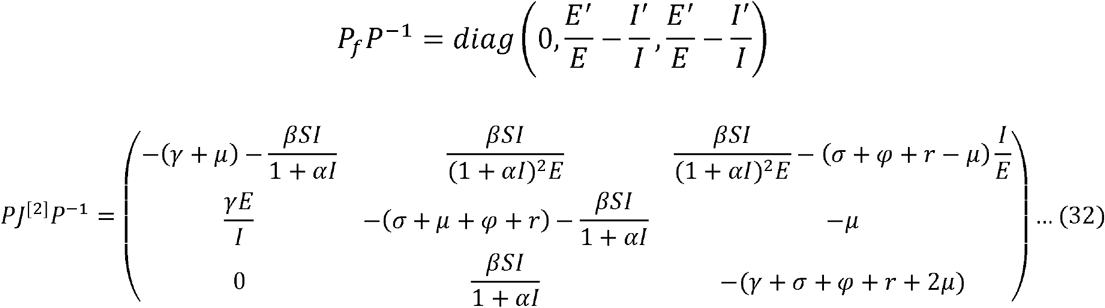

The matrix 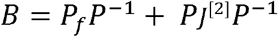 can be written in matrix form as

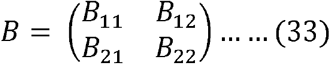

Where

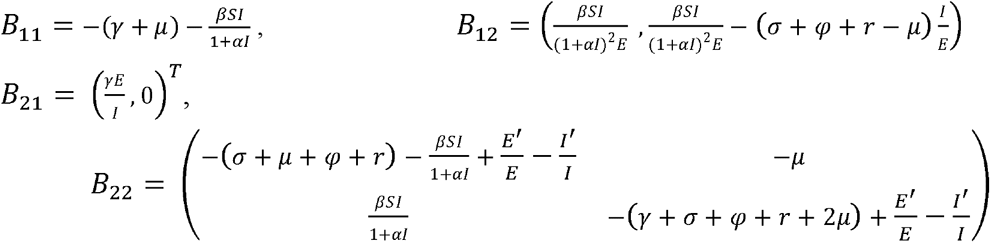

Let (*u, v, w*) be a vector in *ℝ*^3^, its norm ║·║ is defined as

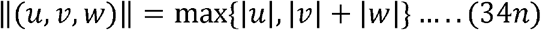

Let *μ*(*B)* be the Lozinskiĭ measure with respect to this norm. Thus, we have

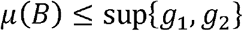

where *g*_1_ = *B*_11_ *+ |B*_12_ *|* and *g*_2_*=μ*_2_(*B*_22_)*+ |B*_21_*|*; *|B*_12_*|* and *|B*_21_*|* are matrix norms with respect to *l*_1_ vector norm and *μ*_2_ denotes the Lozinskiĭ measure with respect to *l*_1_ vector norm.

Then,

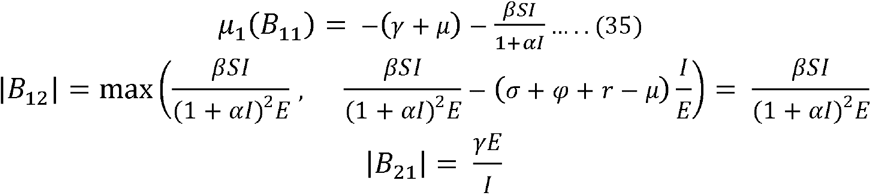

Note that since *B*_11_ is a scalar, its Lozinskiĭ measure with respect to any vector norm *μ*_1_ is equal to *B*_11_. Now calculating *μ*_2_(*B*_22_), taking the non-diagonal elements of each columns of *B*_22_ in absolute value, and then adding to the corresponding columns of the diagonal elements, we have

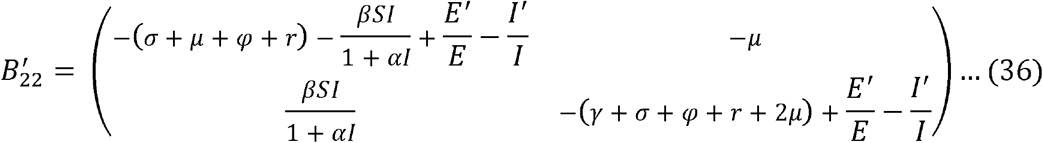

Take the maximum of the two diagonal elements of 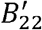, we have

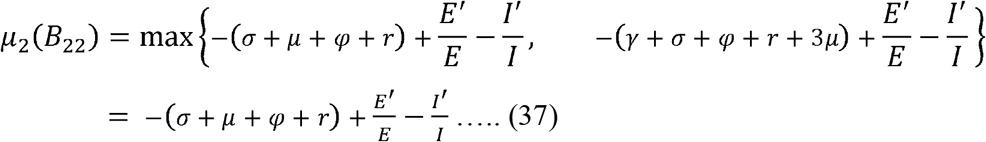

Therefore, we have

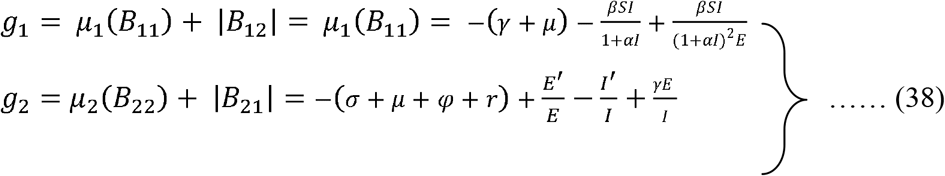

From the system of equations in (5) we have

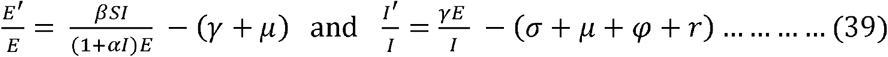

Then we have

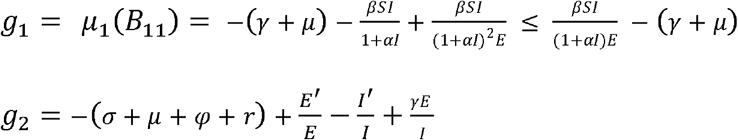

from which we obtain

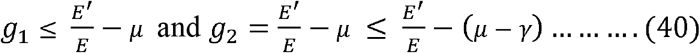

Also, we have

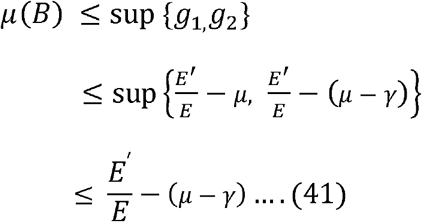

Integrating both sides simultaneously, we have

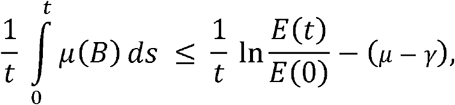

so

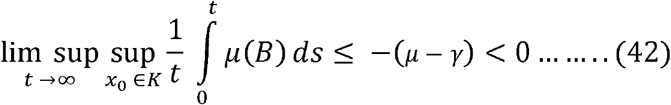

by the hypothesis *μ*−*γ >* 0. Thus, by [31], we have that *P*^*^ is globally asymptotically stable. ■

We will now analyze the global stability of the endemic steady states when *I*^*^ *> I*_0_. After reducing the system of equations in (18) using 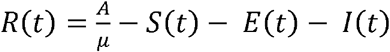, we eliminate the *R*(*t*) component from the first equation of system (18) to obtain a three-dimensional model.

### Theorem 7.3

Let *Q*^*^(*S, E*^*^, *I*^*^, *R*^*^) be an endemic equilibrium of (4) with *I*^*^ *> I*_0_. If *μ* − *γ >* 0 then the endemic equilibrium *Q*^*^ is globally stable.

#### Proof

Since *I*^*^ *> I*_0_, *Q*^*^ is an equilibrium for system (18). The Jacobian matrix of system (18) is given as

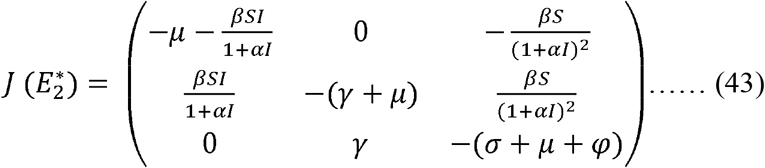

and its second additive matrix is

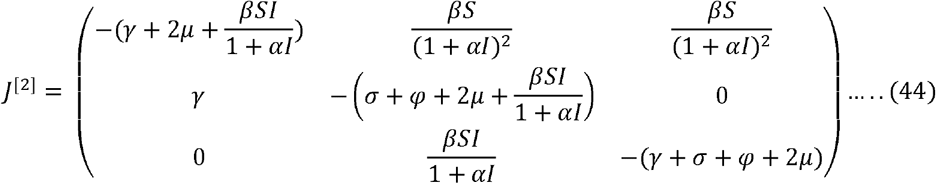

Choose the function 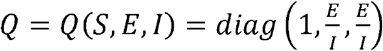; then 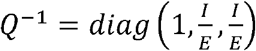 and

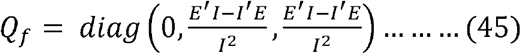

Then we have

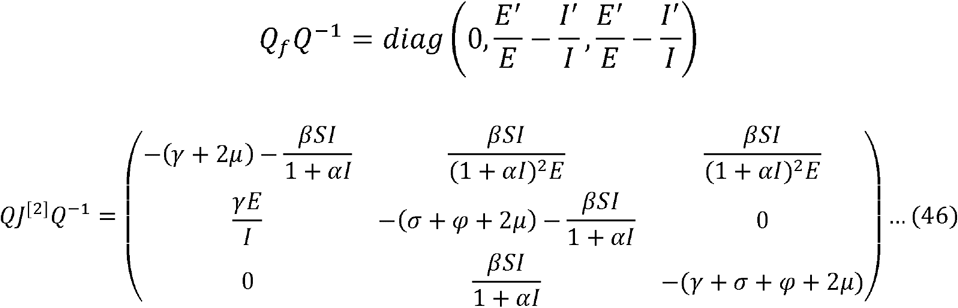

The matrix 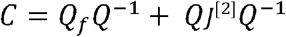 can be written in matrix form as

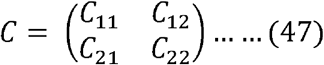

Where

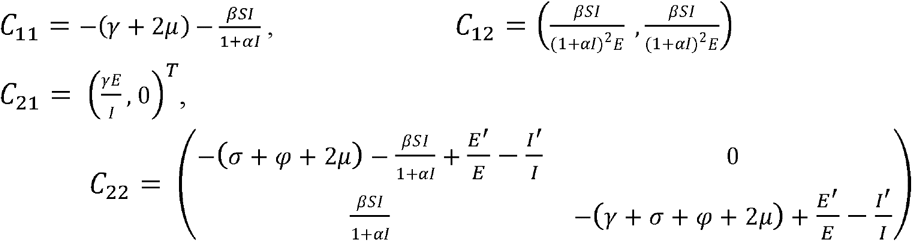

Let (*u, v, w*) be a vector in ℝ^3^, its norm ║·║ is defined as

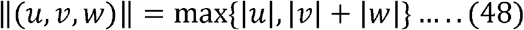

Let *μ*(*C*) be the Lozinskiĭ measure with respect to this norm. Thus, we choose

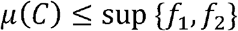

where *f*_2_ = *C*_11_ *+ |C*_12_*|* and *f*_2_ = *μ*_2_(*C*_22_) *+ |C*_21_*|*; *|C*_12_*|* and *|C*_21_*|* are matrix norms with respect to *l*_1_ vector norm and *μ*_2_ denotes the Lozinskiĭ measure with respect to *l*_1_ vector norm.

Then,

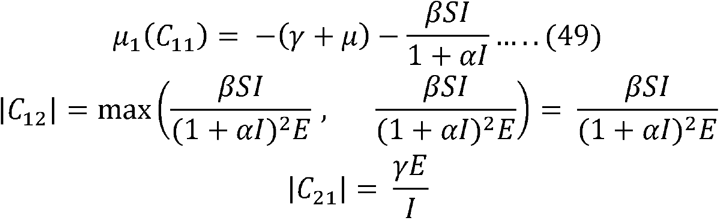

Note that since *C*_11_ is a scalar, its Lozinskiĭ measure with respect to any vector norm *μ*_1_ is equal to *C*_11_. Clearly, to obtain *μ*_2_(*C*_22_), taking the non-diagonal elements of each columns of *C*_22_ in absolute value, and then add it to the corresponding columns of the diagonal elements, we have

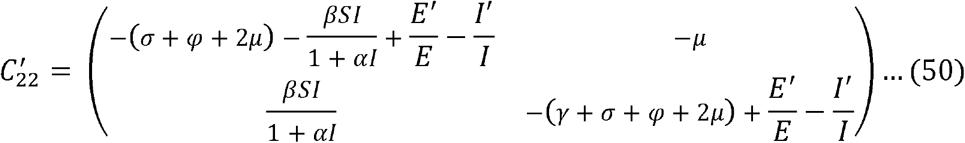

Take the maximum of the two diagonal elements of 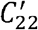, we have

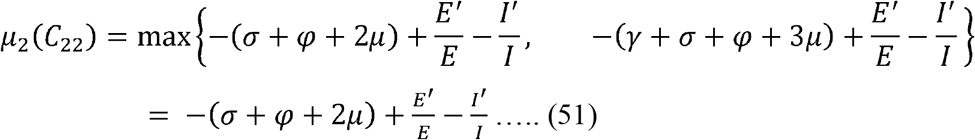

We obtain

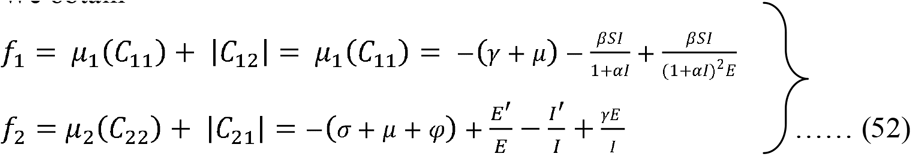

From the system of equations in (18), we have

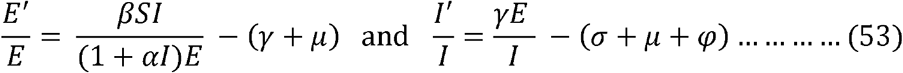

Then we have

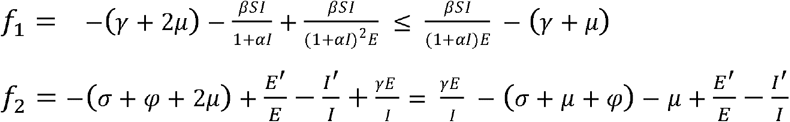

From which we obtain

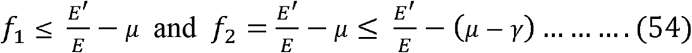

Also, we have

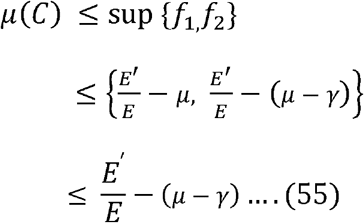

Integrating both sides simultaneously, we have

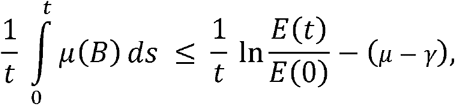

so

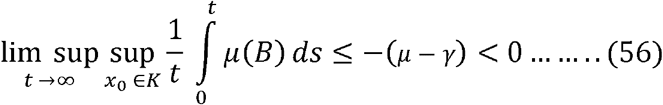

by the hypothesis *μ* − *γ >* 0. Thus, by [31], we have that *Q*^*^ is globally asymptotically stable. ■

## 8. Conclusion

In this paper, we studied the global analysis of an SEIRS epidemic model capturing saturated incidence with treatment response. In the theoretical study of this model, we obtain the basic reproduction number, which explains the dynamic behaviour of the model showing that when *R*_0_ is less than a certain critical value, which we called *R*_1_, the disease-free equilibrium is globally asymptotically stable, that is, the disease dies out. We also determine the existence of the local and global stability of the disease-free and endemic equilibria and found that the efficiency of the treatment response such as contact tracing, quarantine, case search, pool testing, advanced medical technologies, increased trained personnel, funding medical research, etc, and strict adherence to inhibitory measures such as personal hygiene, social distancing, stay-at-home orders, etc, deployed in tackling the COVID-19 pandemic determines whether the disease will die out or become endemic. Hence, how long COVID-19 pandemic stays with us depends on how much we are willing to take responsibility as individuals and government.

## Data Availability

No external source

https://www.researchgate.net/profile/David_Oluyori2

## Notes

### Competing Interest Statement

The authors have declared no competing interest.

### Funding Statement

No funding from any external source

### Author Declarations

IRB approval was not required.

